# Digital Twin-Based Risk Stratification Integrating Aortic Shape and Wall Mechanics in Acute Uncomplicated Type B Aortic Dissection

**DOI:** 10.64898/2026.11.05.26367281

**Authors:** Minliang Liu, Zhongxi Zhou, Yuhang Du, Hannah L. Cebull, Hai Dong, John N. Oshinski, John A. Elefteriades, Rudolph L. Gleason, Bradley G. Leshnower

## Abstract

**Objective:** This study evaluated whether patient-specific digital twins integrating 3D aortic anatomy and structural wall stress could identify patients with acute uncomplicated type B aortic dissection (AUTBAD) at high risk for adverse aortic events.

**Methods:** We analyzed CTAs from 50 patients extracted from a 400 AUTBAD patient database treated with optimal medical therapy from 1999-2025. Digital twin models were constructed, and statistical shape modeling and reduced-order fluid-structure interaction analysis were performed. Anatomic shape and biomechanical stress data were integrated into models predicting freedom from aortic events, defined as intervention or aortic-related death, using leave-one-out cross-validation. A cross-validated risk score was generated for each patient, with higher scores indicating higher event risk.

**Results:** At diagnosis, the median descending aortic diameter was 40.61 mm. During follow-up, 35 patients experienced an aortic event: 28 interventions (56%) and 7 aortic-related deaths (14%). Stratification by descending aortic diameter above or below 40.61 mm yielded higher-versus lower-risk aortic event-free survival of 72.0% versus 84.0% at 1 year and 27.8% versus 56.7% at 6 years (p=0.035; C-index=0.590). In the cross-validated multidimensional model combining morphology and structural stress, higher-versus lower-risk event-free survival was 60.0% versus 96.0% at 1 year and 12.5% versus 68.9% at 6 years (p<0.001; C-index=0.792).

**Conclusions:** Digital twin modeling integrating morphology and wall stress provided greater risk separation than univariable predictors in AUTBAD patients. With independent validation, patient-specific AUTBAD risk stratification using geometric and biomechanical data may ultimately contribute to identification of high-risk patients who may benefit from early endovascular repair.

**Central Picture:** 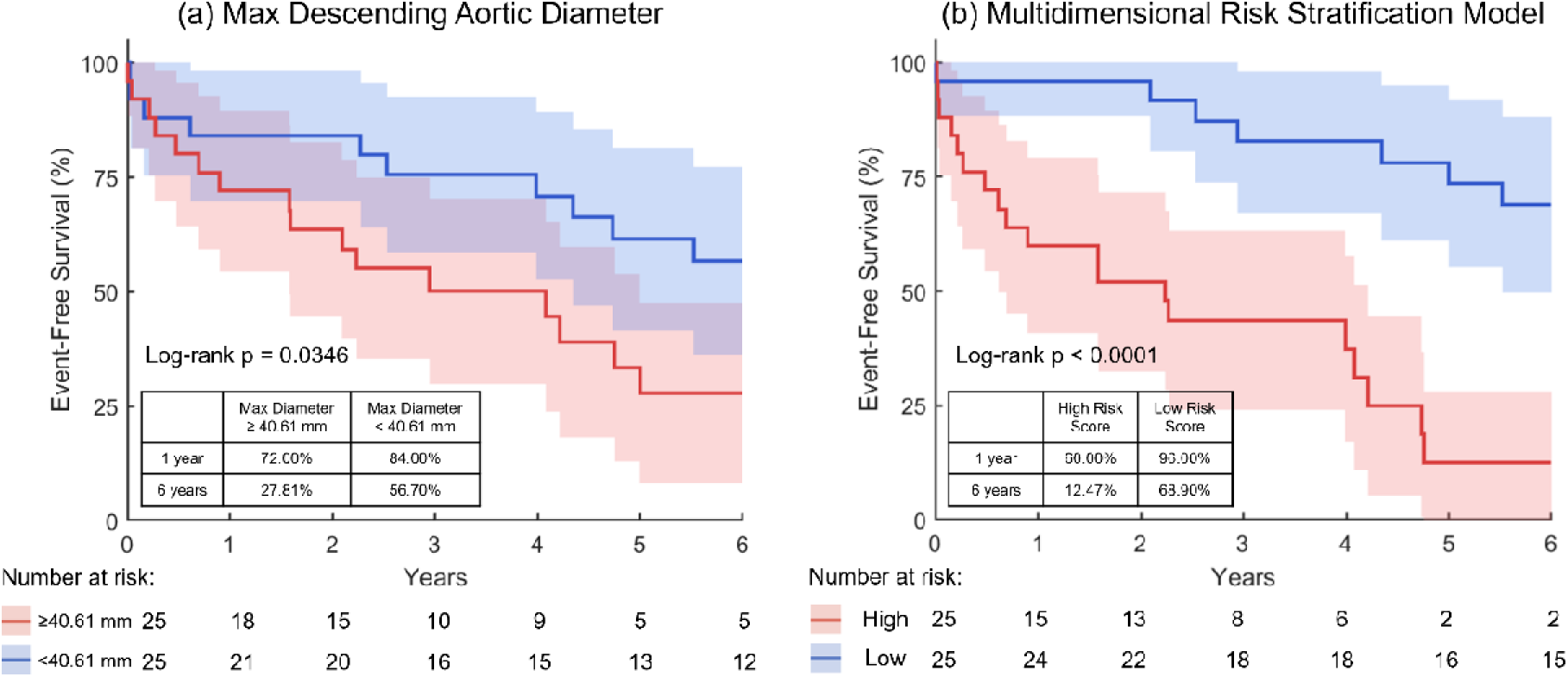

Multidimensional digital twin model stratifies aortic event-free survival in AUTBAD

**Central Message:** Combining patient-specific 3D aortic morphology and wall stress may improve identification of medically managed AUTBAD patients at high risk for aortic intervention or aortic-related death.

**Perspective:** TBAD geometry is highly patient specific, and one-dimensional measurements may not fully reflect patient risk. Digital twin modeling incorporating 3D morphology and wall mechanics may provide a more complete basis for risk stratification. This study demonstrates the feasibility of integrating multidimensional shape and stress data into a risk-stratification model to identify high-risk AUTBAD patients.

## Introduction

The optimal management of acute uncomplicated type B aortic dissection (AUTBAD) remains debated ^1^. Current guidelines recommend optimal medical therapy (OMT), consisting of aggressive blood pressure control with surveillance imaging, as the initial treatment ^1^. Although early survival with OMT exceeds 90%, continued false lumen (FL) pressurization and progressive aortic enlargement contribute to poor long-term outcomes, with 5-year freedom from aortic intervention approximately 50% ^2^. Thoracic endovascular aortic repair (TEVAR) has emerged as an alternative treatment strategy for AUTBAD patients. TEVAR repairs the dissection by implanting an aortic endograft to cover the primary intimal tear. This strategy redirects blood flow into the true lumen (TL), which promotes FL thrombosis and improves aortic remodeling ^3, 4^. TEVAR provides equivalent early survival and may improve midterm outcomes compared with OMT ^3, 4^. However, these benefits must be weighed against cost and associated procedural risks, including stroke, spinal cord ischemia, and retrograde type A dissection ^1, 4^. The ideal treatment strategy for AUTBAD would utilize a patient-specific risk stratification model that could identify those patients at high risk for subsequent aortic enlargement who may benefit from early TEVAR while allowing lower-risk patients to remain on OMT and avoid an unnecessary procedure.

Numerous clinical and anatomic markers have been evaluated as predictors of aortic growth and OMT failure in patients with AUTBAD ^5, 6^. Among these markers, maximum aortic diameter remains the most consistently identified and validated predictor of OMT failure across studies ^5, 7^. However, diameter is a single, one-dimensional measurement that cannot adequately describe the complete 3-dimensional (3D) anatomy of the dissected aorta, which limits its ability to characterize patient-specific risk of aortic expansion ^6, 8^. Furthermore, FL hemodynamics and the biomechanical properties of the false lumen wall have been shown to play integral roles in predicting aortic expansion in TBAD, yet the current understanding of these important factors are still relatively unknown ^9, 10^. These limitations have prompted our group to develop patient-specific digital twin models that incorporate the complete 3D clinical imaging data. Using statistical shape modeling (SSM), we previously quantified the complex 3D anatomy of TBAD and identified shape features associated with subsequent aortic growth and OMT failure ^8^. We have also identified structural wall stress derived from reduced-order fluid-structure interaction (FSI) analysis as a predictor of both the rate and spatial distribution of aortic growth ^11, 12^. Despite their potential value, further insights into aortic remodeling and AUTBAD progression may remain hidden within these highly complex, multidimensional anatomic and biomechanical data. An analytical strategy is therefore needed to uncover these insights and integrate them into a unified risk-stratification model relating patient-specific aortic anatomy and wall mechanics to AUTBAD patient outcomes.

This study evaluated digital twin-derived 3D aortic anatomy and wall mechanics for predicting aortic event-free survival in AUTBAD. SSM-derived shape and FSI-derived stress features were integrated into a regression-based risk-stratification model. We hypothesized that the combined features would better identify patients at high risk for adverse aortic events than individual anatomic or biomechanical predictors.

## Methods

### Study Sample

This retrospective study was approved by the Emory University Institutional Review Board (IRB00109646 and STUDY00002737). The cohort included 50 patients with AUTBAD initially managed with OMT at Emory and Yale Hospitals between 1999 and 2025. Diagnostic contrast-enhanced CTAs of sufficient quality and coverage through the iliac arteries were reviewed sequentially by de-identified study ID to obtain approximately balanced groups with and without iliac involvement (24 and 26 patients, respectively). These diagnostic CTAs were used for FSI and SSM analyses. During follow-up, an aortic event was defined as the first occurrence of open or endovascular aortic intervention or aortic-related death. Indications for intervention were (1) an aortic diameter of ≥5.5 cm, or ≥5.0 cm in patients with a connective tissue disorder; (2) rapid aortic growth of ≥5 mm within 12 months after the first year; or (3) symptomatic aneurysm. Deaths were classified as aortic-related or nonaortic based on medical record review and telephone follow-up conducted by clinical research nurses, with adjudication by the site principal investigators (J.A.E. and B.G.L.).

### Fluid-Structure Interaction Analysis

Diagnostic CTAs were used to construct patient-specific 3D digital twins from the aortic root to the common iliac arteries. Segmentation in 3D Slicer delineated the TL and FL walls, dissection flap, thrombus when present, and branches including the brachiocephalic, left common carotid, left subclavian, celiac, and superior mesenteric arteries. Solid-phase FL thrombus was segmented using the aortic wall’s minimum and maximum Hounsfield unit values as thresholds and modeled as a solid component ^11^. Volumetric FSI meshes were generated in HyperMesh (Altair Engineering Inc.). Figures 1a and 1b show the segmentation and FSI mesh.

**Figure 1.**
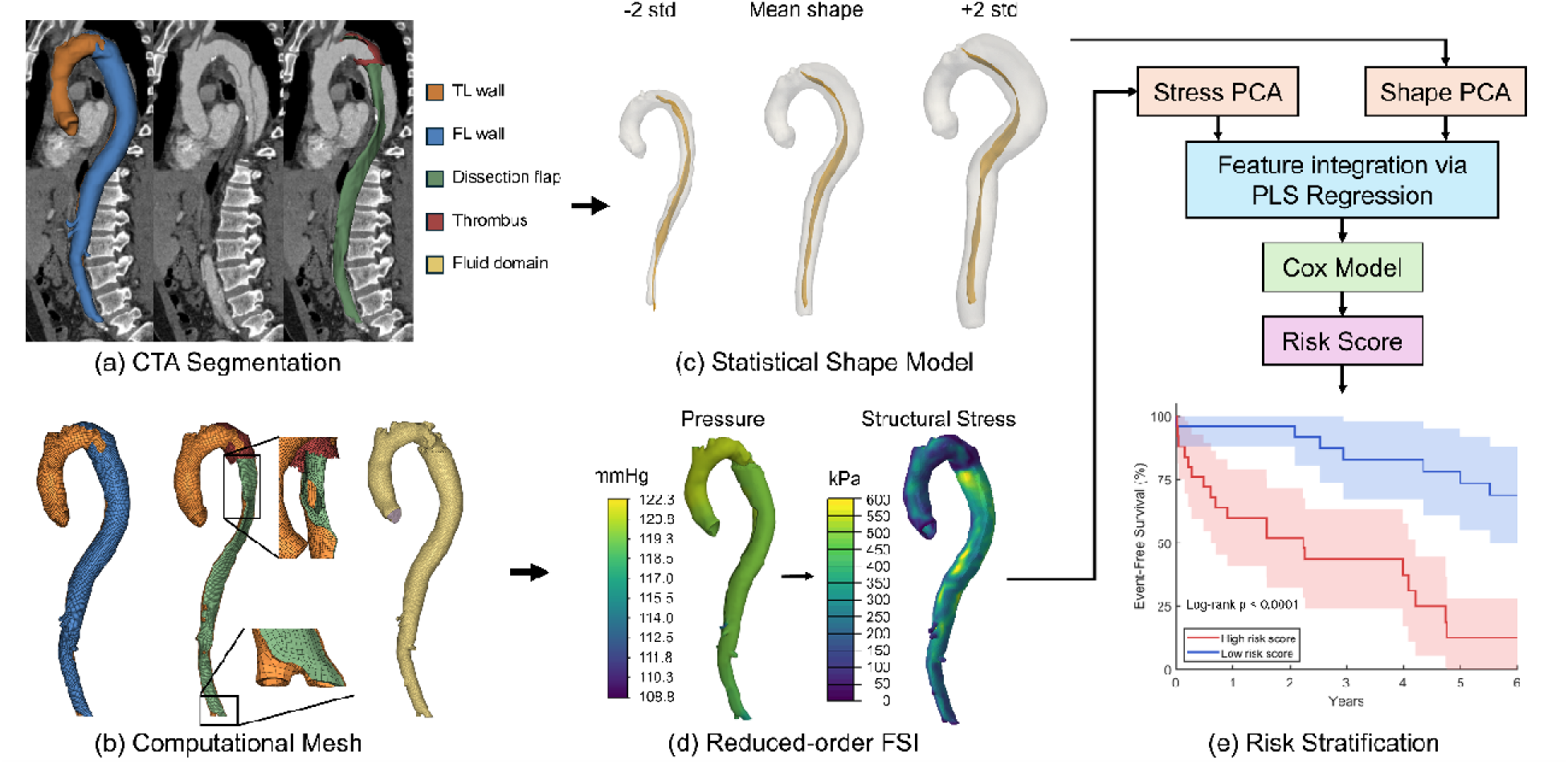
FSI, statistical shape modeling, and PCA-PLS-Cox workflow. (a) Diagnostic CTA segmentation; (b) volumetric FSI mesh; (c) aortic wall and flap shape model; (d) pressure and structural stress; and (e) integration of shape and stress features into individualized risk scores.

Reduced-order FSI simulations calculated structural wall stress for 50 patients using methods previously described and validated by our group ^11, 12^. The analysis consisted of 2 steps. First, blood flow through each patient’s AUTBAD anatomy was modeled using computational fluid dynamics, with peak aortic valve velocity measured by patient-specific echocardiography prescribed as the inlet condition. The calculated pressure was calibrated to the patient’s brachial blood pressure. Second, the pressure distribution obtained from the fluid simulation was applied as the loading condition in finite element analysis to calculate structural wall stress. Structural stress represents mechanical tension within the aortic wall under blood-pressure loading. Higher stress may contribute to adverse remodeling and progressive aortic enlargement.

### Statistical Shape Modeling

A SSM was constructed from the segmented aortic geometries of all 50 patients using our previously described approach ^8^. The SSM uses the complete 3D anatomy, rather than measurements at individual locations, to characterize differences in aortic shape across the cohort. Both the outer aortic wall and dissection flap were included. Entry and re-entry tears, FL thrombus, and patency were not explicitly represented in the SSM. To allow direct comparison of 3D anatomy across patients, each patient’s aortic wall and dissection flap were represented using standardized templates containing 10317 and 1989 points, respectively. Each template point corresponded to the same anatomic region in every patient, which allowed local differences in aortic and flap shape to be compared across different patients. Principal component analysis (PCA), a standard approach for feature extraction from high-dimensional data, was then used to characterize the dominant variations in aortic shape across patients, which yielded 49 PCA shape features. Figure 1c illustrates the first shape feature, which represents the largest overall differences in dissected aortic anatomy among patients in the study population.

### Risk Stratification and Statistical Analysis

Structural stress distributions computed by FSI and shape features extracted by SSM were used to develop a multidimensional model for predicting aortic event-free survival. PCA summarized thousands of spatial variables as a compact representation of variation across patients. For mean-centered data, the maximum number of nonzero modes was one fewer than the number of patients used to fit PCA.

Within the reduced PCA representation, bootstrap-based feature selection used variable importance in projection (VIP >1) ^13^ as the criterion. Features selected in at least 90% of bootstrap samples were retained. Partial least squares (PLS) regression used null-Cox deviance residuals as its response (PLSDR) ^14^, which incorporated event status and the estimated cumulative hazard at the observed event or censoring time. PLS integrated the selected shape and stress PCA modes into a single feature that served as the sole predictor in a Cox proportional hazards model. This PCA reduction and PLS integration helped contain model complexity in this small cohort. The Cox model generated individualized risk scores, with higher scores indicating greater aortic event risk.

Model performance was evaluated using leave-one-out cross-validation (LOOCV). In each of 50 iterations, PCA-PLS-Cox fitting and feature selection used only 49 patients to predict the held-out patient’s risk score. The 50 cross-validated scores were split at their median to define higher- and lower-risk groups.

For comparison, patients were also stratified using individual predictors, including maximum descending aortic diameter, mean structural wall stress of the descending aorta, and iliac artery involvement. Diameter and stress were dichotomized at their median values. Parsimonious Cox models using continuous diameter, diameter plus iliac involvement, and diameter plus mean structural wall stress were also evaluated using LOOCV.

Aortic event-free survival was estimated using Kaplan-Meier analysis and compared using log-rank tests. Predictive performance was assessed from continuous Cox risk scores and predicted survival probabilities using Harrell’s C-index, time-dependent AUC, and Brier scores, with 95% confidence intervals estimated and the stability of predictive performance assessed using a delete-one-patient analysis. A p value < .05 was considered statistically significant.

Regional stress-growth rate associations were assessed using linear mixed-effects regression ^12^. The average patient-specific Pearson correlation was tested against r = 0.2.

## Results

### Patient Demographics and Comorbidities

Table 1 summarizes the demographic, comorbidity, and anatomic characteristics of the 50 patients. The median age was 58 years, and 5 patients (10%) had a connective tissue disorder. During follow-up, 35 patients experienced a first aortic event: 28 interventions and 7 aortic-related deaths before intervention. One additional aortic-related death occurred after intervention (Table E3). Events comprised 3 acute (≤14 days), 1 subacute (15-90 days), and 4 chronic (>90 days) aortic-related deaths, and 2 subacute and 26 chronic interventions (Tables E4 and E5). On the index CT scan, the median maximum descending aortic diameter was 40.6 mm, and the primary intimal tear was located in Ishimaru zone 3 in 25 patients (50%). Median time to the first aortic event or last follow-up was 3.88 years (IQR, 1.58-7.63 years; Table 1). Four patients (8%) were lost to follow-up and censored at their last event-free encounter. The distribution of diagnosis years is shown in Figure E1.

**Table 1.**
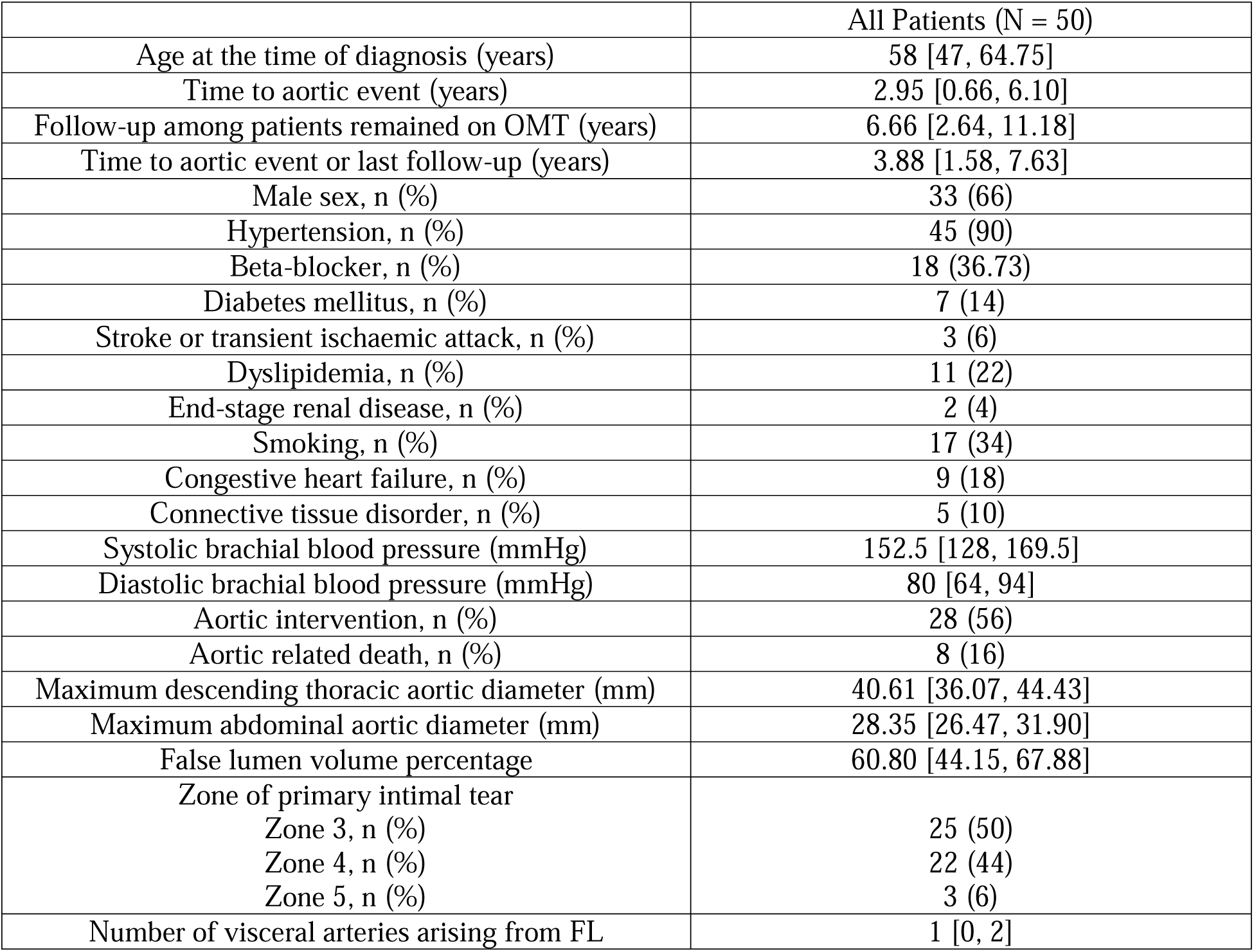
Demographic, clinical, and anatomic characteristics of the cohort. Visceral artery count includes the celiac, superior mesenteric, inferior mesenteric, and renal arteries. Data are median (25th, 75th percentiles) or number (percentage). Missing values were excluded.

### Risk Stratification Using Individual Predictors and Parsimonious Cox Models

Maximum descending aortic diameter and spatially averaged structural wall stress, both derived from geometries reconstructed from the initial diagnostic CTA, were dichotomized at their median values of 40.61 mm and 181.02 kPa, respectively, whereas iliac artery involvement was classified as present or absent. Patients with a maximum descending aortic diameter <40.61 mm had significantly higher aortic event-free survival than those with a diameter ≥40.61 mm at 1 year (84.00% vs 72.00%) and 6 years (56.70% vs 27.81%; log-rank p=.0346; Figure 2a). Patients with structural wall stress <181.02 kPa had higher event-free survival than those with stress≥181.02 kPa at 1 year (84.00% vs 72.00%) and 6 years (60.44% vs 24.88%; log-rank p=.0170; Figure 2b). Patients with iliac artery involvement also had higher event-free survival than those without iliac involvement at 1 year (91.67% vs 65.39%) and 6 years (55.66% vs 31.21%; log-rank p=.0306; Figure 2c). LOOCV C-index, time-dependent AUC, and Brier scores with 95% confidence intervals are reported in Table E1. C-indices for the parsimonious Cox models were 0.657 for diameter, 0.627 for diameter plus iliac involvement, and 0.678 for diameter plus stress.

**Figure 2.**
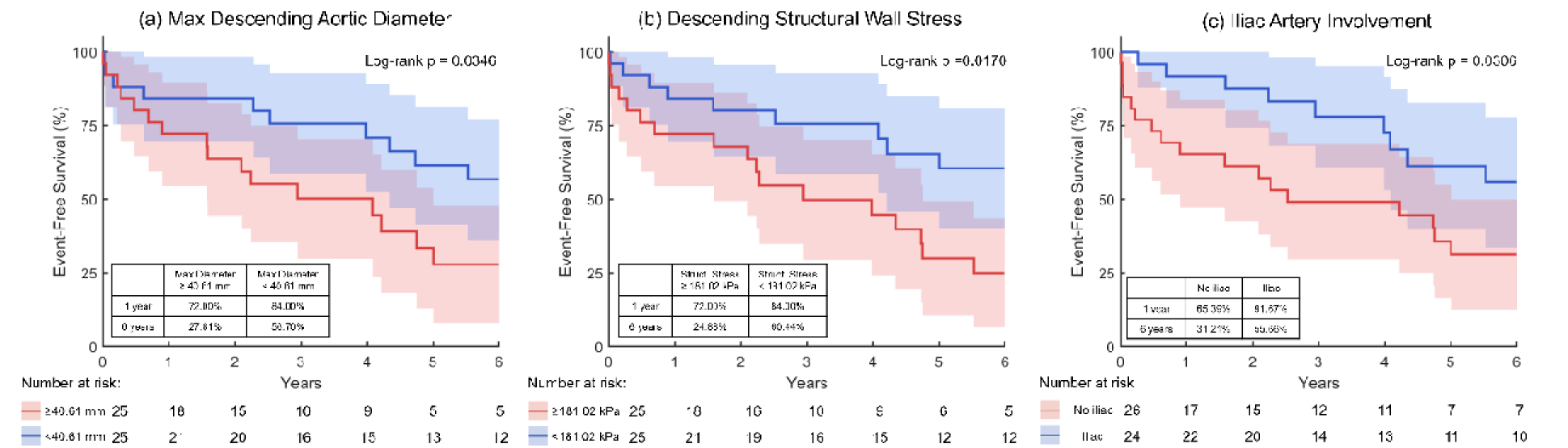
Kaplan-Meier estimates of aortic event-free survival stratified by (a) maximum descending aortic diameter, (b) mean descending structural wall stress, and (c) iliac artery involvement. Shading represents 95% confidence intervals.

### Risk Stratification Using the Multidimensional PCA-PLS-Cox Model

Three PCA-PLS-Cox models used stress, shape, or combined features. For the structural stress-only model, the higher-risk group had significantly lower aortic event-free survival than the lower-risk group at 1 year (64.00% vs 92.00%) and 6 years (21.67% vs 66.56%; log-rank p=.0012; Figure 3a). Similar separation was observed with the shape-only model at 1 year (64.00% vs 92.00%) and 6 years (12.47% vs 69.17%; log-rank p<.0001; Figure 3b). For the combined stress and shape model, event-free survival was 60.00% versus 96.00% at 1 year and 12.47% versus 68.90% at 6 years in the higher- and lower-risk groups, respectively (log-rank p<.0001; Figure 3c). LOOCV C-indices were 0.691, 0.737, and 0.754 for the stress, shape, and combined models, respectively. Predictive performance was relatively stable across delete-one-patient analyses (Table E2).

**Figure 3.**
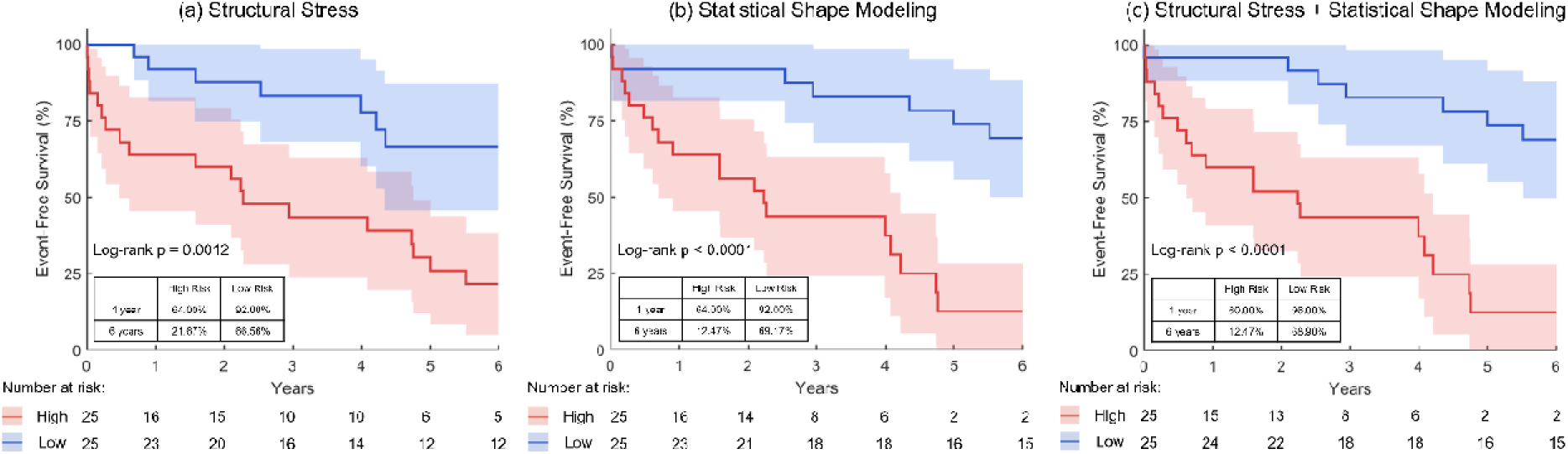
Kaplan-Meier estimates of aortic event-free survival stratified by PCA-PLS-Cox models using (a) stress, (b) shape, and (c) combined stress and shape features. Shading represents 95% confidence intervals.

The PLS-derived composite shape and structural stress features are shown in Figures 4 and 5. The higher-risk direction of the shape feature was characterized by a larger aortic diameter, longer centerline, and shorter flap, consistent with limited distal dissection and absence of iliac involvement (Figure 4). The stress feature showed greater proximal descending aortic stress in the higher-risk direction and greater distal descending aortic stress in the lower-risk direction (Figure 5).

**Figure 4.**
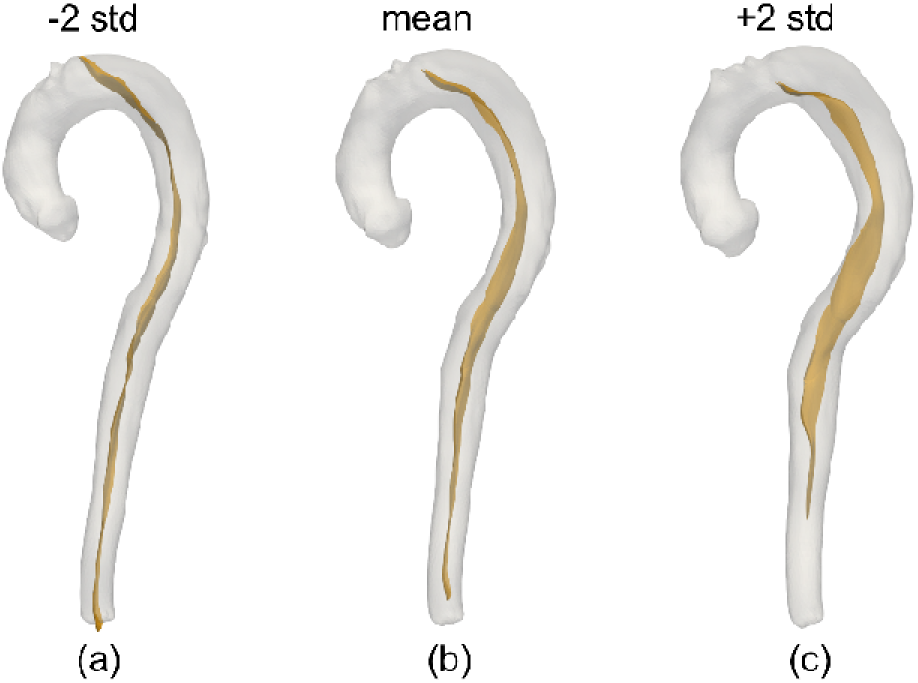
PLS-derived composite shape feature on the mean dissected aorta. “+2 std” and “−2 std” show the mean shape shifted 2 standard deviations toward higher and lower risk, respectively.

**Figure 5.**
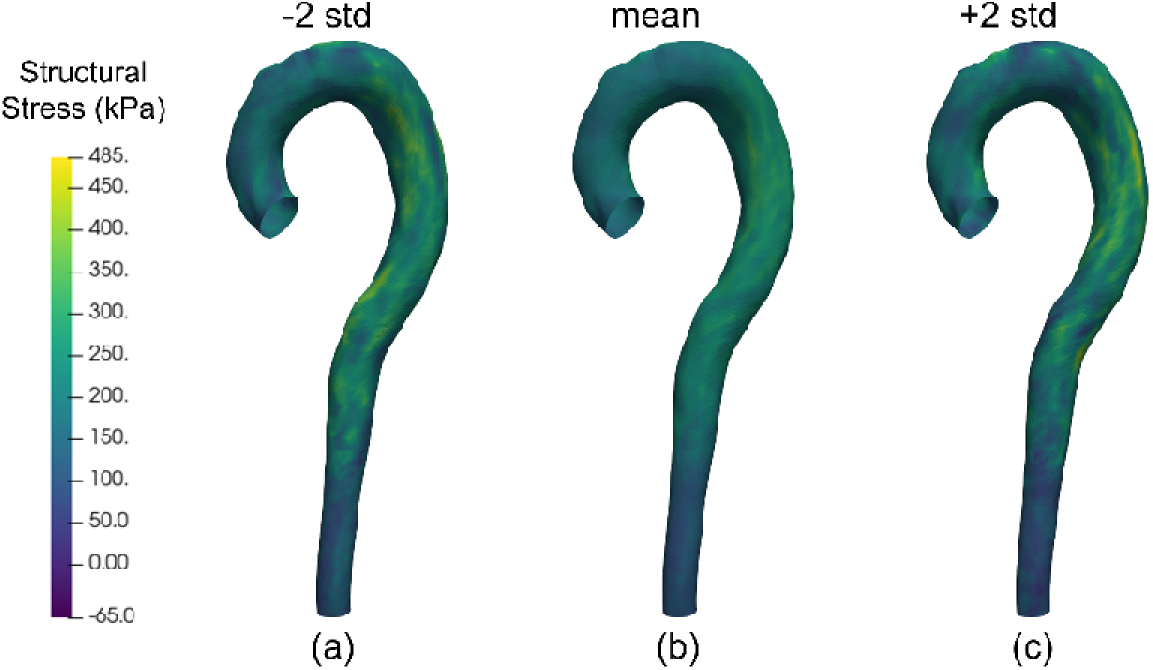
PLS-derived composite structural stress feature on the mean dissected aorta. “+2 std” and “−2 std” show the mean stress field shifted 2 standard deviations toward higher and lower risk, respectively.

Structural stress exhibited a significant positive association with aortic growth rate (fixed-effect p = 0.0003). A one-sided test of patient-specific correlations showed that the association exceeded the weak-correlation threshold of R = 0.2 (p = 0.0127).

## Discussion

The current controversy regarding the treatment of patients with AUTBAD is whether TEVAR should replace OMT as primary therapy. Advocates for OMT maintain that TEVAR should be reserved for TBAD complicated by malperfusion or rupture, while TEVAR supporters contend that all TBAD’s are complicated and should undergo endovascular therapy. An alternative approach is to perform selective TEVAR in the acute phase on patients who are at high risk for false lumen aneurysm formation and treat the remaining lower risk patients with OMT alone. This strategy would avoid the potential risks and adverse effects associated with implanting aortic endografts in patients who are unlikely to experience significant aortic growth. These potential benefits must be weighed against the risks of TEVAR, including stroke, spinal cord ischemia, and retrograde type A dissection. In the current study we present our initial work towards the development of the 1^st^ patient-specific risk stratification model for patients with AUTBAD using 3D anatomic and biomechanical information extracted from patient-specific digital twin models developed from standard clinical imaging.

Previous investigations to define anatomic predictors of OMT failure in TBAD have been largely unsuccessful, and maximum aortic diameter remains as the sole predictor of OMT failure that has been consistently identified and validated across multiple studies ^5^. However, diameter alone does not capture the complete 3D anatomy and wall mechanics of the dissected aorta. We therefore incorporated complementary anatomic and biomechanical information into our risk model. SSM characterized variation in 3D aortic geometry across the cohort, and reduced-order FSI quantified structural wall stress throughout the TL and FL. These 3D data were integrated into a compact representation of aortic anatomy and structural stress. The multidimensional model provided greater separation between higher- and lower-risk groups than individual predictors or parsimonious Cox models.

Patient-specific geometry in TBAD may limit the ability of any single measurement to accurately characterize patient risk. No individual measurement is likely to capture the complete hemodynamic and mechanical environment of the FL, which depends on its inflow/outflow balance. FL fluid dynamics and wall stress reflect the impact of the complex 3D aortic geometry incorporated into the model, including tear location, size and configuration, dissection extent, FL and TL size, and visceral and intercostal arteries arising from the FL ^6^. Visualization of the PLS-derived composite shape and stress features (Figs 4, 5) further illustrated that risk reflected anatomic and biomechanical characteristics throughout the dissected aorta rather than any single local measurement.

Our preliminary analysis suggested that a larger aortic diameter, longer centerline, and limited dissection extent (flap ending proximal to the aorto-iliac bifurcation) were associated with higher aortic-event risk. These findings are consistent with prior work identifying aortic diameter and aortic centerline length as predictors of adverse events and OMT failure in AUTBAD ^7, 15^. They are also consistent with our previous finding that dissections without iliac artery involvement have increased aortic-event risk ^16^, which may reflect greater FL pressurization. Interestingly, our analysis showed that higher structural wall stress in the distal descending aorta was associated with a lower risk of aortic events. This association may also reflect differences in the spatial distribution of FL pressurization. Lower stress in the proximal descending aorta accompanied by higher stress distally may indicate proximal FL thrombosis or a more distal primary intimal tear, either of which could reduce FL pressure and promote more favorable aortic remodeling ^17, 18^.

This study has several limitations. First, the cohort was relatively small (n = 50), in part because the FSI modeling pipeline required substantial manual effort to process the complex 3D geometries derived from clinical imaging. Given the small cohort, model complexity, and absence of independent validation, the findings should be interpreted with caution (see the ROADMAP study ^19^). The small cohort also limited separate analyses by disease phase and event type. Future work will incorporate deep learning-based segmentation and meshing methods ^20^ to automate the modeling process and facilitate validation in larger cohorts with independent test datasets. Such automation could also support the development of software for potential application in clinical settings. Second, the composite endpoint combines aortic-related death and intervention. A larger initial aortic diameter could contribute to earlier attainment of criteria for intervention. Third, although medical therapy remained the standard initial treatment for AUTBAD and formal intervention criteria were unchanged, changes in imaging and clinical practice over the prolonged study period may have introduced variability. These temporal effects warrant further adjustment in larger cohorts. Fourth, the FSI models assumed regionally uniform wall thicknesses based on experimental data ^10^ and represented only the solid phase of thrombus. Patient-specific wall thickness and biphasic thrombus warrant further investigation using higher-resolution CTA and more sophisticated models.

## Conclusions

This proof-of-concept digital twin framework demonstrated the feasibility of integrating patient-specific 3D aortic anatomy and structural wall stress to stratify aortic event-free survival in AUTBAD. The multidimensional PCA-PLS-Cox model provided greater separation between higher- and lower-risk patients than individual anatomic and biomechanical predictors. The composite features suggested that FL aneurysmal degeneration reflects geometric and biomechanical characteristics throughout the dissected aorta. With independent validation in larger cohorts and prospective studies, this framework may ultimately contribute to the identification of high-risk patients who may benefit from early TEVAR.

## IRB

STUDY00002737. Approved July 23, 2021. Need for consent was waived.

## Data Availability

All data produced in the present study are available upon reasonable request to the corresponding author

## Acknowledgements

The authors thank Kristina Porte for significant contributions to clinical data acquisition.

## Glossary of Abbreviations

AUTBAD: acute uncomplicated type B aortic dissection
TBAD: type B aortic dissection
CTA: computed tomography angiography
OMT: optimal medical therapy
TEVAR: thoracic endovascular aortic repair
TL: true lumen
FL: false lumen
FSI: fluid structure interaction

---

**Figure E1.**
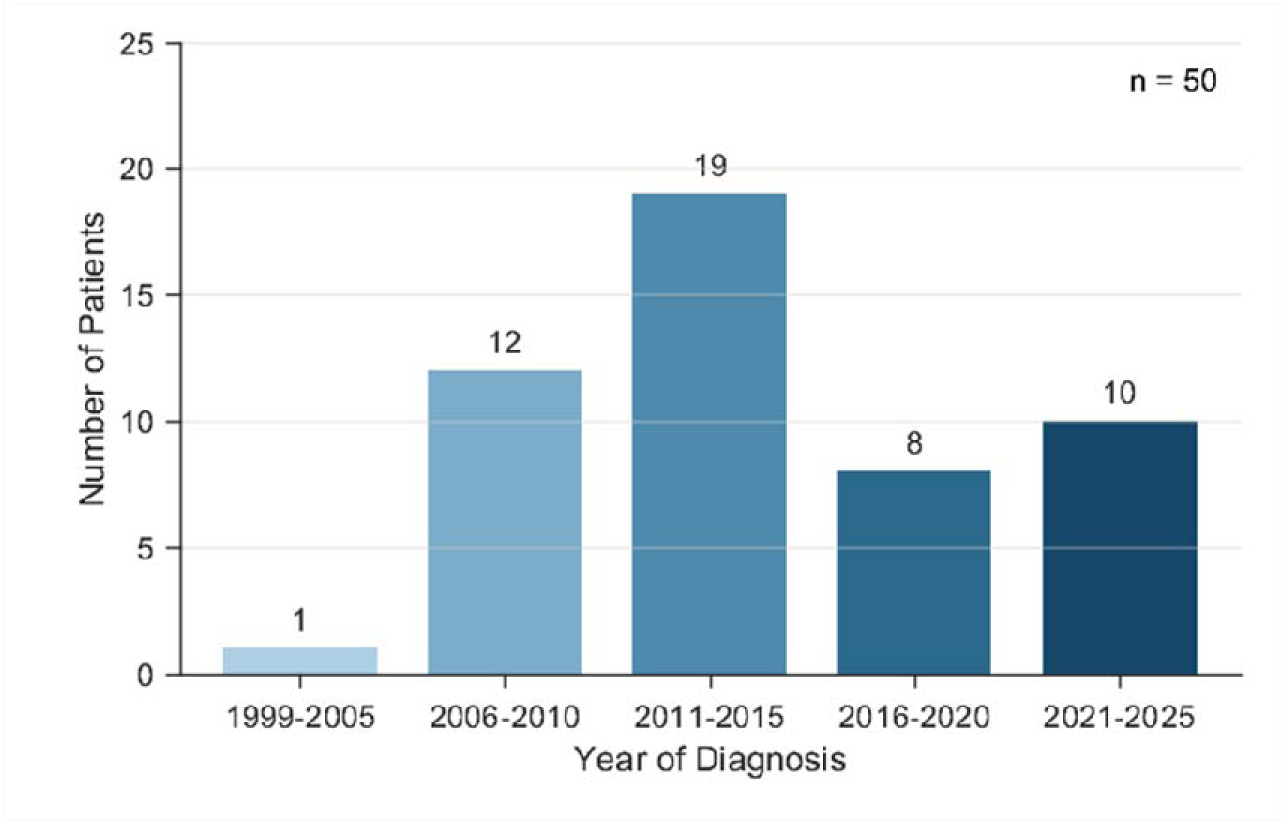
Temporal distribution of AUTBAD diagnosis in the study cohort.

**Figure E2.**
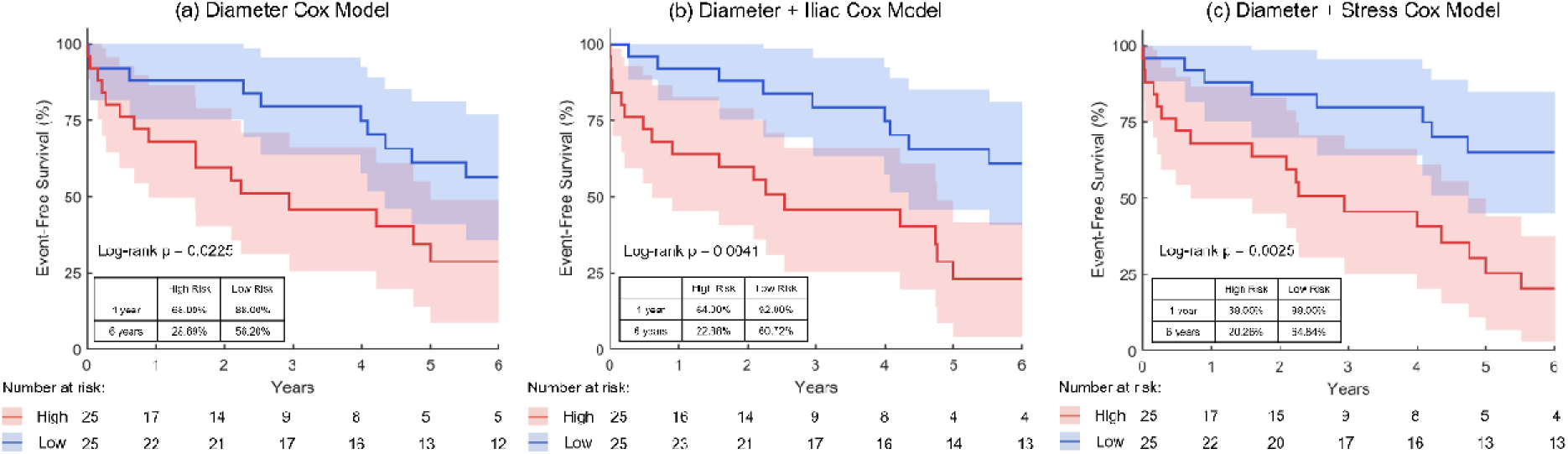
Kaplan-Meier estimates of aortic event-free survival stratified by parsimonious Cox models using (a) continuous maximum descending aortic diameter, (b) continuous maximum descending aortic diameter and iliac artery involvement, and (c) continuous maximum descending aortic diameter and mean descending structural wall stress. Shading represents 95% confidence intervals.

**Figure E3.**
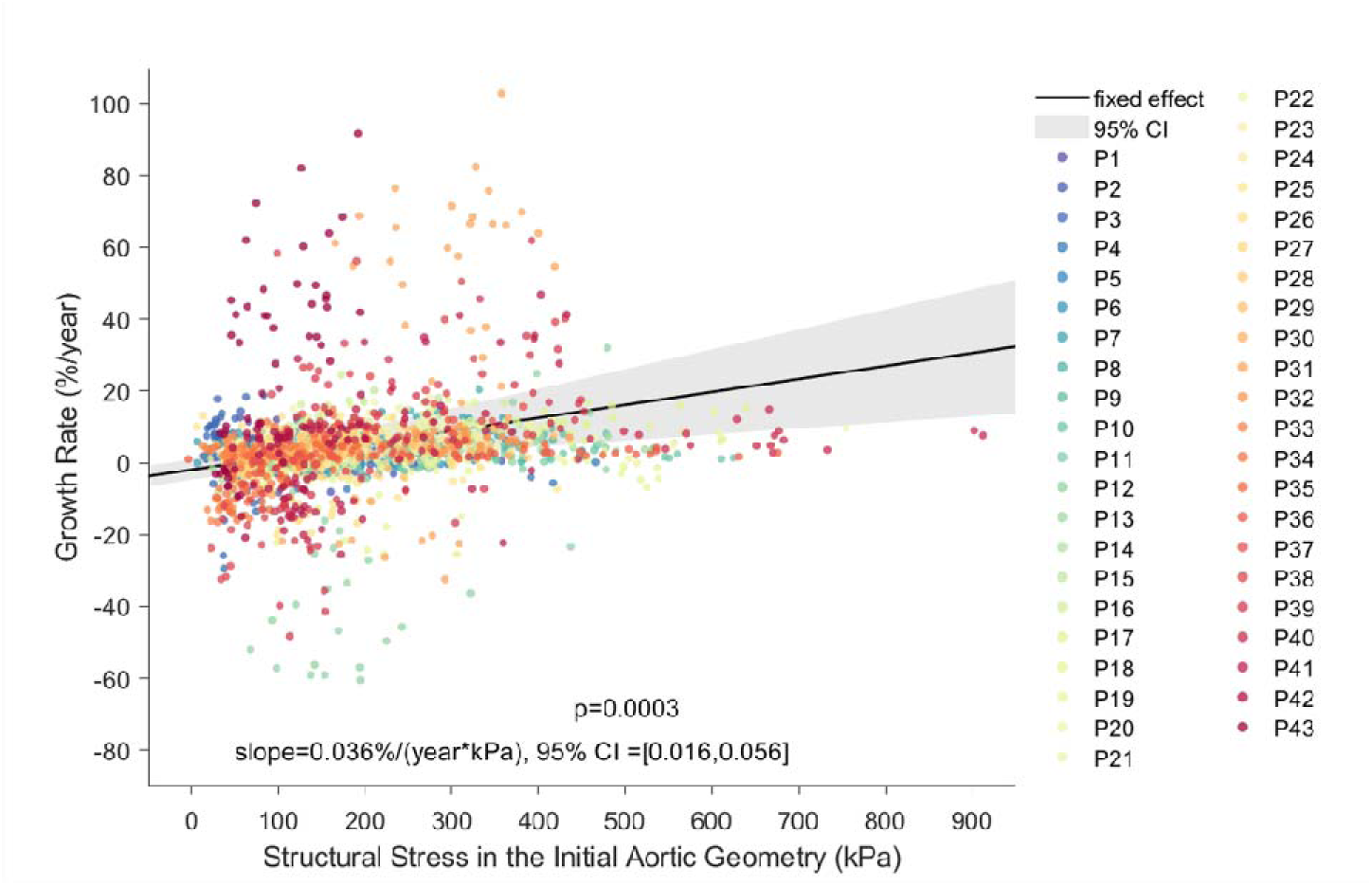
Linear mixed-effects regression analysis demonstrates a significant positive fixed-effect association between structural stress in the initial aortic geometry and aortic growth rate (p=0.0003; slope=0.036%/(year*kPa)).

**Table E1.** Predictive performance of individual predictors, parsimonious, and PCA-PLS-Cox models. 95% confidence intervals are presented in the brackets.

| Predictor / model |  | Harrell’s C-index | Time-dependent AUC |  | Time-dependent Brier score |  |
|---|---|---|---|---|---|---|
|  |  |  | 1 year | 6 years | 1 year | 6 years |
| Individual predictors (no cross-validation) | Diameter ≥ 40.61mm | 0.590 [0.461, 0.719] | 0.557 [0.349, 0.765] | 0.692 [0.526, 0.857] | 0.174 [0.107, 0.240] | 0.229 [0.190, 0.269] |
|  | Stress ≥ 181.02kPa | 0.560 [0.442, 0.678] | 0.531 [0.337, 0.726] | 0.655 [0.476, 0.834] | 0.173 [0.106, 0.241] | 0.234 [0.200, 0.268] |
|  | Iliac involvement | 0.623 [0.530, 0.717] | 0.691 [0.544, 0.838] | 0.614 [0.458, 0.771] | 0.157 [0.099, 0.215] | 0.233 [0.191, 0.275] |
| Parsimonious Cox models (LOOCV) | Diameter | 0.657 [0.473, 0.841] | 0.671 [0.452, 0.891] | 0.792 [0.570, 1.000] | 0.188 [0.115, 0.262] | 0.244 [0.199, 0.288] |
|  | Diameter + iliac | 0.627 [0.465, 0.789] | 0.674 [0.474, 0.873] | 0.702 [0.500, 0.904] | 0.175 [0.108, 0.242] | 0.245 [0.183, 0.307] |
|  | Diameter + stress | 0.678 [0.564, 0.792] | 0.667 [0.527, 0.806] | 0.842 [0.702, 0.982] | 0.192 [0.116, 0.269] | 0.243 [0.184, 0.303] |
| PCA-PLS-Cox models (LOOCV) | Stress | 0.691 [0.498, 0.885] | 0.739 [0.407, 1.000] | 0.761 [0.458, 1.000] | 0.149 [0.046, 0.253] | 0.211 [0.061, 0.360] |
|  | Shape | 0.737 [0.621, 0.853] | 0.839 [0.693, 0.984] | 0.826 [0.666, 0.986] | 0.152 [0.066, 0.237] | 0.179 [0.072, 0.286] |
|  | Shape + stress | 0.754 [0.598, 0.910] | 0.839 [0.687, 0.991] | 0.841 [0.640, 1.000] | 0.157 [0.067, 0.247] | 0.163 [0.023, 0.302] |

**Table E2.** Standard deviations of model performance in the delete-one sensitivity analysis.

| PCA-PLS-Cox model | Harrell’s C-index | Time-dependent AUC |  | Time-dependent Brier score |  |
|---|---|---|---|---|---|
|  |  | 1 year | 6 years | 1 year | 6 years |
| Stress | 0.014 | 0.024 | 0.022 | 0.007 | 0.011 |
| Shape | 0.009 | 0.011 | 0.012 | 0.006 | 0.008 |
| Shape + stress | 0.011 | 0.011 | 0.015 | 0.007 | 0.010 |

**Table E3.** First aortic events during overall follow-up and the 6-year analysis period.

| First aortic event | Overall follow-up, n (%) | 6-year analysis period, n (%) |
|---|---|---|
| Aortic intervention | 28 (56) | 21 (42) |
| Aortic-related death | 7 (14) | 5 (10) |
| Total first composite events | 35 (70) | 26 (52) |

**Table E4.** Aortic interventions in patients undergoing optimal medical therapy. M, Male; F, female.

| Sex | Days from diagnosis to intervention | Indication for intervention |
|---|---|---|
| M | 94 | Rapid growth |
| M | 254 | Rapid growth |
| M | 580 | Aneurysm |
| F | 818 | Aneurysm |
| M | 1076 | Aneurysm |
| M | 1457 | Aneurysm |
| M | 1490 | Aneurysm |
| M | 1582 | Aneurysm |
| M | 2014 | Aneurysm |
| M | 2440 | Aneurysm |
| F | 2767 | Aneurysm |
| M | 3830 | Aneurysm |
| M | 4036 | Aneurysm |
| M | 51 | Rapid growth & Aneurysm |
| F | 77 | Rapid growth |
| M | 173 | Rapid growth & Aneurysm |
| M | 329 | Rapid growth & Aneurysm |
| F | 576 | Aneurysm |
| F | 588 | Rapid growth |
| M | 761 | Aneurysm |
| M | 831 | Aneurysm |
| M | 925 | Aneurysm |
| M | 1469 | Aneurysm |
| M | 1726 | Aneurysm |
| M | 1855 | Aneurysm |
| M | 2514 | Aneurysm |
| M | 2836 | Aneurysm |
| F | 4816 | Aneurysm |

**Table E5.**
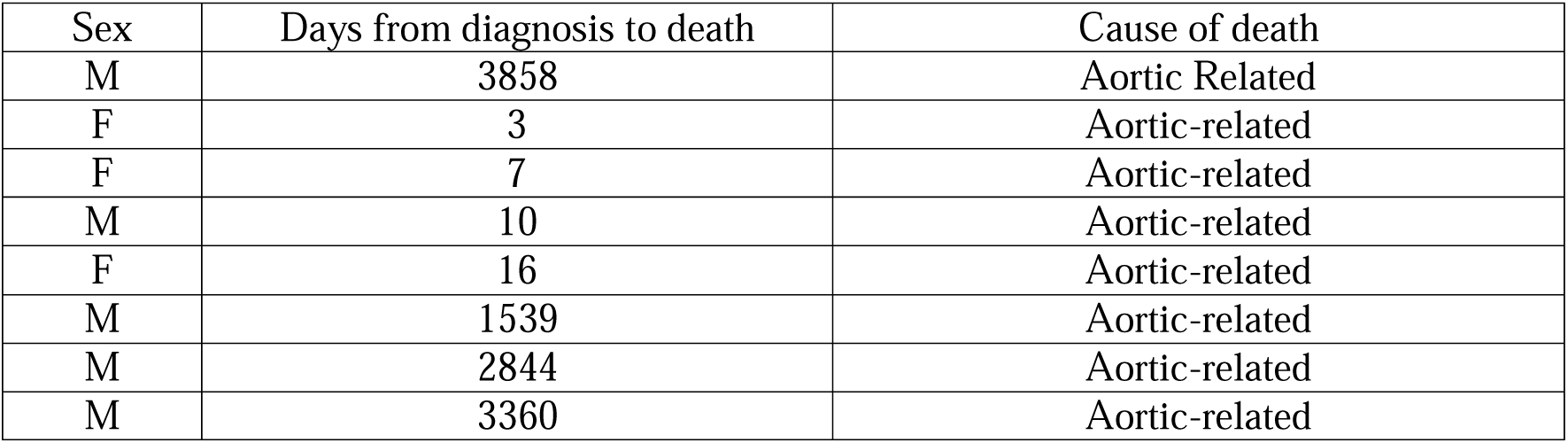
Aortic-related deaths in patients undergoing optimal medical therapy. M, Male; F, female.

## References

1. MacGillivray TE, Gleason TG, Patel HJ, et al. The Society of Thoracic Surgeons/American Association for Thoracic Surgery clinical practice guidelines on the management of type B aortic dissection. The Journal of Thoracic and Cardiovascular Surgery. 2022;163:1231–1249.

2. Lou X, Chen EP, Duwayri YM, et al. The impact of thoracic endovascular aortic repair on long-term survival in type B aortic dissection. The Annals of thoracic surgery. 2018;105:31–38.

3. Nienaber CA, Kische S, Rousseau H, et al. Endovascular repair of type B aortic dissection: long-term results of the randomized investigation of stent grafts in aortic dissection trial. Circulation: Cardiovascular Interventions. 2013;6:407–416.

4. Brunkwall J, Kasprzak P, Verhoeven E, et al. Endovascular repair of acute uncomplicated aortic type B dissection promotes aortic remodelling: 1 year results of the ADSORB trial. European Journal of Vascular and Endovascular Surgery. 2014;48:285–291.

5. Spinelli D, Benedetto F, Donato R, et al. Current evidence in predictors of aortic growth and events in acute type B aortic dissection. Journal of vascular surgery. 2018;68:1925–1935.

6. Sailer AM, Van Kuijk SM, Nelemans PJ, et al. Computed tomography imaging features in acute uncomplicated Stanford type-B aortic dissection predict late adverse events. Circulation: Cardiovascular Imaging. 2017;10.

7. Lou X, Duwayri YM, Chen EP, et al. Predictors of failure of medical management in uncomplicated type B aortic dissection. The Annals of thoracic surgery. 2019;107:493–498.

8. Liu M, Dong H, Mazlout A, et al. The role of anatomic shape features in the prognosis of uncomplicated type B aortic dissection initially treated with optimal medical therapy. Computers in biology and medicine. 2024;170:108041.

9. Cebull HL, Liu M, Piccinelli M, et al. The use of 4-dimensional flow magnetic resonance imaging and fluid structure interaction analysis to predict failure of medical therapy in acute uncomplicated type B aortic dissection. JTCVS techniques. 2024;27:40–44.

10. Dong H, Liu M, Cebull HL, et al. Protective biomechanical and histological changes in the false lumen wall in chronic type B aortic dissection. JTCVS open. 2025;23:60–68.

11. Liu M, Du Y, Cebull HL, et al. Investigating the role of structural wall stress in aortic growth prognosis in acute uncomplicated type B aortic dissection. Biomechanics and Modeling in Mechanobiology. 2026;25:11.

12. Du Y, Wu Y, Cebull HL, et al. Structural Stress as a Predictor of the Rate and Spatial Location of Aortic Growth in Uncomplicated Type B Aortic Dissection. IEEE Transactions on Biomedical Engineering. 2026.

13. Chong I-G, Jun C-H. Performance of some variable selection methods when multicollinearity is present. Chemometrics and intelligent laboratory systems. 2005;78:103–112.

14. Bastien P. Deviance residuals based PLS regression for censored data in high dimensional setting. Chemometrics and Intelligent Laboratory Systems. 2008;91:78–86.

15. Liang X, Dong H, Schmid M-PH, et al. Changes in aortic centerline length and curvature predict aortic diameter growth in chronic type B aortic dissection. JTCVS open. 2026:101680.

16. Huckaby LV, Dong H, Liu M, et al. Iliac artery involvement determines optimal therapy in acute uncomplicated type B aortic dissection. JTCVS Structural and Endovascular. 2026:100110.

17. Girish A, Padala M, Kalra K, et al. The Impact of Intimal Tear Location and Partial False Lumen Thrombosis in Acute Type B AorticáDissection. The Annals of thoracic surgery. 2016;102:1925–1932.

18. Parker LP, Reutersberg B, Syed MB, et al. Proximal false lumen thrombosis is associated with low false lumen pressure and fewer complications in type B aortic dissection. Journal of Vascular Surgery. 2022;75:1181–1190.

19. Fleischmann D, Mastrodicasa D, Willemink MJ, et al. Predicting late adverse events in uncomplicated Stanford Type B aortic dissection: results from the ROADMAP validation study. Circulation: Cardiovascular Imaging. 2025;18.

20. Qian L, Chen J, Gong R, Sun W, Liu M, Liang L. Shape deformation networks for automated aortic valve finite element meshing from 3D CT images. Medical Imaging 2026: Clinical and Biomedical Imaging2026:13929: 13424–13433.

